# Does genetic risk for depression accelerate biological aging?

**DOI:** 10.1101/2025.07.23.25328503

**Authors:** Darina Kirilina

## Abstract

Major Depressive Disorder is a major contributor to aging-related health burdens. Preventing and managing depression symptoms are suggested strategies to reduce these burdens. However, the cooccurrence of depression and aging-related disease/disability could arise from reverse causation (i.e. aging-->depression) or confounding by a common cause (aging<- ->depression). We investigated the connection between depression and biological aging, a root cause of aging-related health decline, using US Health and Retirement Study (HRS) data and a genetic design to address potential confounding. We tested associations among polygenic scores measuring genetic risk for depression, longitudinal measurements of depression symptoms, and a measurement of biological aging in 3,806 White and 1,207 Black HRS participants. Results showed that participants with higher depression polygenic scores and more depression symptoms were biologically older than those of the same chronological age with lower genetic risk and fewer symptoms. However, depression symptoms mediated only a fraction of the association between genetic risk and biological aging. Findings suggest that the link between depression and aging burden may not be fully addressable through depression treatment in later life. Further research is needed to determine whether our findings reflect the accumulated effects of depressive symptoms earlier in life or some other pathway linking genetic risk for depression with accelerated biological aging.

## INTRODUCTION

Major Depressive Disorder (MDD) is one of the major contributors to overall disease burden, including conditions like diabetes, heart disease, cognitive decline, Alzheimer’s disease^1^, and premature death^2^. Older adults are at an increased risk for experiencing depression^3^. World Health Organization reports that approximately 5.7% of adults aged 60 years and older experiences depression^4^. In USA around 7 million American older adults experience depression each year^5^.

Biological aging involves the gradual loss of integrity and resilience in cells, tissues, and organs over time. Depression has been linked to biological irregularities that contribute to accelerated biological aging^6^. A meta-analysis by Ridout et al. found a significant negative association between depression and telomere length^7^. Additionally, depression impairs mitochondrial function, crucial for energy production and stress adaptation^8^, and metabolic dysregulation, characterized by high glucose and triglycerides and low high-density lipoprotein (HDL) levels, which increase the risk of psychiatric disorders and further contribute to the acceleration of biological aging^9^. Moreover, individuals with MDD often exhibit chronic inflammation, another key driver of aging-related processes^10^.

This link between depression and aging burden suggests that treating depression in the older adults might help alleviate the accelerated biological aging, improving the overall health and the quality of life of older individuals. The Global Burden of Disease study emphasizes the potential of mental health interventions to reduce the overall disease burden, particularly in aging populations, by lowering Disability-Adjusted Life Years (DALYs) and improving life expectancy^2^. Epidemiological studies further underscore that early detection and treatment of depression can significantly improve physical health outcomes and decrease mortality rates, especially in individuals with chronic diseases such as diabetes^11^.

However, epidemiological associations may overestimate healthspan benefits of later-life depression treatment and prevention due to potential confounding pathways. One concern is reverse causation, in which aging-related health declines contribute to the development of depression. In addition to poor health contributing to depressed mood, mechanisms of biological aging may independently contribute to the heightened risk of depression and anxiety disorders among older adults^7,8,9^. For example, vascular depression theory proposes that cerebrovascular damage and the subsequent alterations in brain structure and function can increase the likelihood of experiencing depressive symptoms^14^. A second concern are common causes, including overlapping biological mechanisms that could drive both depression and accelerated aging. For example, the body’s stress response plays a key role in both depression and aging^15^. Similarly, oxidative stress can reduce essential brain-supporting factors, linking depression with faster aging^15^. Finally, depression-aging associations could reflect effects established early in life. Individuals with poor mental and cognitive health in their younger years are at heightened risk of developing age-related diseases^16^ and show signs of accelerated functional decline even before the onset of age-related diseases^17^.

Depression differs in its episodes and severity among individuals, making it difficult to capture its full trajectory with a single measurement. Genetic analysis can play a crucial role in understanding the bidirectional relationship between accelerated biological aging and depression risk. Genetic predisposition to depression remains stable throughout the lifespan, unlike the dynamic factors influenced by aging, such as physiological changes^18^. The research indicates that the genetic profiles can be considered reliable predictors of depression and anxiety risk across different age groups, from middle-aged to elderly populations^19–21^. Also, recent studies have emphasized the associations between genetics and biological age, particularly focusing on PhenoAge, one of the widely applied biological age measures^22^.

Polygenic risk scores (PRS) have emerged as an significant area of research, offering a way to measure genetic susceptibility to diseases^23^. A key advantage of PRS is that they can be used for their ability to predict diseases and adjust for genetic background without requiring a detailed genetic model ^24,25,26^. However, some research caution that PRS can introduce bias in certain contexts^26^. The predictive accuracy of a PRS is affected by factors like genome-wide association studies (GWAS) sample size, trait heritability, and the consistency of genetic architecture across environments^27^. Nonetheless, PRS can still serve as useful markers for genetic predispositions related to factors, associated with biological aging, such as hormonal imbalances, inflammatory processes, and biological dysfunction^28,29^.

We investigated the relationship between PRS for MDD and biological aging, with a focus on understanding how these associations are influenced by depression symptoms observed in later life. Using data from 3,806 White and 1,207 Black participants in the US Health and Retirement Study (HRS), we tested whether higher genetic risk for depression and a greater burden of depressive symptoms were associated with accelerated biological aging. We also examined whether the predictive power of polygenic scores differed between older adults of European and African ancestry. Finally, we analyzed whether depression symptoms mediated the association between genetic risk and biological aging.

## MATERIALS AND METHODS

### Sample

This study is based on data from participants in the Health and Retirement Study (HRS) who provided blood-chemistry data as part of the Venous Blood Study (VBS), as well as genetic data obtained from the HRS Polygenic Genetic Data.

The HRS is a publicly accessible national panel survey that targets individuals aged 50 and above, as well as their spouses^30^. The HRS has been conducted biennially since 1992, with a new cohort of 51-to 56-year-olds and their spouses enrolled every six years to maintain national representativeness. The questionnaire covers a wide range of topics including health, cognition, family, employment, disability, and income. The total response rates across all waves of the HRS range from 74% to 93%^31^. The phenotype and demographic data were sourced from the RAND HRS Longitudinal File 2020 (V1) provided by the research organization RAND^32^. The RAND HRS File includes fifteen waves of Core Interview data across seventeen survey years. As of the most recent data release in March 2023, the RAND HRS File comprised data from 42,406 individuals in 26,595 households ^33^.

The VBS, conducted in 2016 to understand how social factors impact health on a biological level^34^, involved the collection of biomarker data, including blood-chemistry data, from a subset of 9,286 HRS participants who consented to venous blood draws^35^.

Genetic risk measures, specifically polygenic risk scores, were derived from the HRS Polygenic Scores - Release 4.3 2006-2012 Genetic Data (Feb 2021). Starting in 2006, the HRS introduced an Enhanced Face-to-Face Interview (EFTF) involving physical assessments, body measurements, blood and saliva sample collection, and a self-administered survey covering psychosocial subjects. About half of households with eligible participants participated in this expanded interview. Polygenic scores for depression, based on the 2018 GWAS by the Psychiatric Genomics Consortium’s MDD working group, were used for European ancestry (N=12,090) and African American ancestry (N=3,100) groups separately.

In this study the HRS data curated by the RAND Corporation was linked with the data collected as part of the HRS’s 2016 VBS and the data obtained from the HRS Polygenic Genetic Data. The final sample was divided into two separate datasets for the participants of European (hereafter “White”) (N=3806) and African American (hereafter “Black”) ancestry (N=1207).

## Measures

### Genetic Variables

This study included two types of genetic variables: genetic principal components used to control for ancestry-related confounding in genetic analysis, and polygenic scores used to measure genetic risk for depressive symptoms.

### Principal components

Principal components (PCs) were derived from genome-wide SNP data to identify and adjust for population stratification effects. In this study PCs were incorporated as covariates in the statistical model to account for any potential confounding that could arise from genetic ancestry differences within the study populations.

### Polygenic Scores

Polygenic Score is an index that combines information from millions of single-nucleotide polymorphisms (SNPs) identified across the genome, capturing the cumulative genetic influence on a specific phenotype^36^. Polygenic scores are derived from genome-wide association studies (GWAS), which identify genetic markers (usually SNPs) statistically linked to the trait of interest^37^. This study uses the HRS depression polygenic score obtained from the HRS Polygenic Scores Release 4.3 (from February 2021), a database containing computed polygenic scores for a variety of phenotypes for HRS respondents who provided salivary DNA between 2006 and 2012. The Polygenic Risk Scores (PRS) in this study were calculated using the standardized residuals adjusted for PCs.

### Depressive Symptoms

The HRS collected data on participants’ depressive symptoms using the Center for Epidemiologic Studies Depression (CESD) scale, developed by Laurie Radloff in 1977. This self-report questionnaire includes 20 items measuring various aspects of depression experienced within the past week ^38^, rated on a scale from 0 (rarely or none of the time) to 3 (most or all the time). It assesses feelings such as sadness, loneliness, lack of motivation, and pessimism.

CESD data collection began with the initiation of the HRS in 1992, capturing responses from the initial cohort born between 1931 and 1941. Subsequently, CESD data were collected biennially in alignment with the HRS Core Interviews. This study uses the database “RAND HRS Longitudinal File 2020 (V1)”, which spans fifteen waves of Core Interview data from 1992 to 2020 and includes information from multiple entry cohorts, providing comprehensive longitudinal insights into depressive symptomatology across diverse demographic groups.

### Biological Age

There is no universally recognized definitive measure for biological aging ^39^. There are many proposed methods focusing on different biological levels of analysis. Current state-of-the-art methods use machine-learning methods to sift large numbers of candidate biomarkers and parameterize algorithms that predict aging-related parameters, including chronological age, mortality risk, and rate of decline in system integrity. Algorithms are developed in reference data and then applied in new data to test hypotheses. In our analysis, we focused on the blood-chemistry biological age measure “PhenoAge” originally proposed by Levine^40^. The PhenoAge was developed using elastic-net regression ^41^ to develop a predictive model of survival in data from the US National Health and Nutrition Examination Survey (NHANES) based on chronological age and a panel of 42 clinical laboratory parameters. The resulting algorithm consists of age and nine blood analytes.

For our analysis we selected PhenoAge based on prior validation evidence establishing it as best-in-class among measures of biological age that can be computed from standard clinical laboratory data ^42–45^ and evidence that it is predictive of morbidity, disability, and mortality in Black and White adults in the HRS ^46^. PhenoAge was computed for HRS participants based on Levine’s original algorithm using the BioAge R Package ^43^ applied to blood chemistry data collected by HRS VBS. PhenoAge was calculated for n=9,286 participants. In this study PhenoAge was residualized on chronological age (i.e. we regressed computed PhenoAge on chronological age used the residual values for analysis). We refer to these residuals following convention in the field as “Phenotypic Age Acceleration” (PAA).

Additional variables encoding demographic information were sourced from the RAND HRS File.

## Statistical analysis

All analyses included Black and White participants with complete data on age, gender, CESD scores, PRS, and PAA. Participants with missing data were excluded from the analysis. To estimate the genetic effect of PRS on CESD and PAA and to explore the relationship between CESD and PAA, the multivariate linear regression models were employed, estimated using ordinary least squares regression. The calculations were performed separately for White and Black population groups. All models were adjusted for age and gender.

Model 1 contains the analysis of the association between PRS and CESD across multiple years (1994–2020) to evaluate the genetic effect of PRS on CESD. Model 2 focuses on examining the relationship between genetic predisposition to depression, measured by PRS, and PAA.

To explore the impact of CESD on PAA, we conducted two sets of analyses. The first set used CESD data from 2016, while the second set used the cross-time average CESD score up to 2016. These models helped us understand the influence of depression (as measured by CESD) on PAA (Model 3 and Model 4). Additionally, we tested the combined effects of PRS and CESD on PAA. We included both the PRS and CESD (from 2016 or as a cross-time average) in the regression models to determine their joint influence on phenotypic age acceleration (Model 5 and Model 6).

Additionally, mediation analysis was conducted to explore the role of CESD as a potential mediator in the relationship between the exposure variable (PRS) and the outcome variable (PAA), utilizing a regression-based approach as described by VanderWeele and Vansteelandt ^47^. This method enabled the decomposition of the total effect of the exposure on the outcome into direct and indirect pathways. We specified two regression models for each relationship of interest: one for PAA as a function of PRS, CESD, and covariates, and another for CESD as a function of PRS and covariates (age and gender). We utilized the R package “CMAverse” to estimate direct and indirect effects based on these regressions ^48^.

All analyses were performed in R Studio 2022.07.2.

## RESULTS

We analyzed data for HRS participants with available genetic, depressive-symptom, and biological age data (n=5013). We conducted separate analyses in individuals identifying as White (N=3806) and Black (N=1207) following the approach proposed by HRS in their construction of ancestry-specific principal components. Table 1 represents the final White and Black population summary including the CESD score, PAA data, and the PRS score, divided by Wave Year of interview (1994-2020).

**Table 1.** Demographic Characteristics of the Black and White Population Across Wave Years 1994-2020, Health and Retirement Study Abbreviations: SD, Standard Deviation; CESD, Center for Epidemiologic Studies Depression; PRS, polygenic risk scores; PAA, Phenotypic Age Acceleration. The table represents the final White and Black population summary including the CESD score, PAA data, and the PRS score, divided by Wave Year of interview (1994-2020).

| Wave Year | Sample | Percent of Males (%) | Age, Median (SD) | CESD Score, Median (SD) | PRS Score, Median (SD) | PAA, Median (SD) | Sample | Percent of Males (%) | Age, Median (SD) | CESD Score, Median (SD) | PRS Score, Median (SD) | PAA, Median (SD) |
| --- | --- | --- | --- | --- | --- | --- | --- | --- | --- | --- | --- | --- |
| Black Population |  |  |  |  |  |  | White Population |  |  |  |  |  |
| 1994 | 285 | 31 | 55.71 (5.73) | 1.53 (2.09) | -0.04 (0.95) | 3.31 (9.09) | 1,358 | 51 | 57.68 (4.44) | 0.78 (1.46) | -0.03 (1.01) | 1.56 (8.87) |
| 1996 | 275 | 29 | 57.64 (5.77) | 1.41 (1.83) | -0.07 (0.96) | 3.08 (8.90) | 1,376 | 51 | 59.71 (4.41) | 0.76 (1.38) | -0.03 (1.01) | 1.52 (8.84) |
| 1998 | 401 | 30 | 58.09 (7.02) | 1.91 (2.06) | -0.02 (1.03) | 2.86 (9.31) | 2,092 | 51 | 59.84 (6.39) | 1.05 (1.57) | -0.04 (1.00) | 1.34 (8.79) |
| 2000 | 405 | 31 | 59.87 (7.21) | 1.78 (1.92) | -0.01 (1.02) | 3.11 (9.33) | 2,089 | 51 | 61.87 (6.38) | 1.02 (1.54) | -0.05 (0.99) | 1.29 (8.77) |
| 2002 | 413 | 31 | 61.91 (7.26) | 1.91 (2.12) | -0.01 (1.02) | 3.15 (9.24) | 2,112 | 51 | 63.79 (6.38) | 0.95 (1.59) | -0.05 (1.00) | 1.31 (8.81) |
| 2004 | 601 | 33 | 60.65 (8.30) | 1.84 (2.14) | 0.01 (1.02) | 2.94 (9.47) | 2,768 | 50 | 62.61 (8.22) | 0.96 (1.59) | -0.04 (0.99) | 0.80 (8.68) |
| 2006 | 615 | 33 | 62.45 (8.49) | 1.78 (2.09) | 0.00 (1.01) | 2.95 (9.48) | 2,826 | 51 | 64.60 (8.20) | 1.03 (1.69) | -0.03 (0.99) | 0.78 (8.65) |
| 2008 | 622 | 34 | 64.57 (8.29) | 1.64 (2.02) | 0.00 (1.01) | 3.03 (9.45) | 2,838 | 51 | 66.56 (8.23) | 0.96 (1.60) | -0.03 (1.00) | 0.77 (8.65) |
| 2010 | 1,216 | 35 | 60.83 (9.23) | 1.86 (2.10) | -0.02 (1.00) | 2.68 (9.43) | 3,748 | 51 | 64.83 (9.86) | 1.02 (1.68) | -0.02 (1.00) | 0.26 (8.48) |
| 2012 | 1,224 | 35 | 62.73 (9.25) | 1.82 (2.10) | -0.02 (1.00) | 2.63 (9.48) | 3,766 | 51 | 66.80 (9.88) | 1.05 (1.69) | -0.02 (1.00) | 0.26 (8.48) |
| 2014 | 1,222 | 35 | 64.77 (9.27) | 1.78 (2.09) | -0.02 (1.00) | 2.66 (9.47) | 3,763 | 51 | 68.80 (9.91) | 1.10 (1.74) | -0.02 (1.00) | 0.27 (8.48) |
| 2016 | 1,237 | 35 | 66.76 (9.26) | 1.74 (2.00) | -0.02 (1.00) | 2.68 (9.50) | 3,806 | 51 | 70.79 (9.91) | 1.10 (1.74) | -0.02 (1.00) | 0.27 (8.49) |
| 2018 | 1,083 | 34 | 68.36 (8.78) | 1.69 (1.99) | -0.01 (0.98) | 2.68 (9.41) | 3,360 | 51 | 72.43 (9.59) | 1.17 (1.79) | -0.03 (1.00) | -0.07 (8.27) |
| 2020 | 987 | 32 | 69.67 (8.49) | 1.68 (2.00) | -0.02 (0.99) | 2.15 (9.10) | 3,035 | 51 | 73.49 (9.30) | 1.23 (1.86) | -0.04 (1.00) | -0.51 (7.98) |

### The associations between CESD and PRS

Table 2 represents the associations between the depression score (CESD) and the polygenic risk score (PRS) for each the wave year from 1994 to 2020 for Black and White population (Model 1). For the White population dataset, the analysis revealed consistently positive associations between CESD and PRS across all years. Specifically, the regression coefficient β_PRS_ranged from 0.10 to 0.20 (95% CI: 0.03 to 0.25). These associations were statistically significant at the *P* < 0.001 level for all wave years, indicating a robust relationship between PRS and CESD in the White population over time. In contrast, for the Black population dataset, the associations between CESD and PRS varied across wave years and were generally weaker compared to the White population (range –0.12, 010 (95% CI: −0.39 to 0.35)) and not statistically significant for all wave years. It is important to note that PRS was standardized to Z-score units, while CESD was not standardized.

**Table 2.**
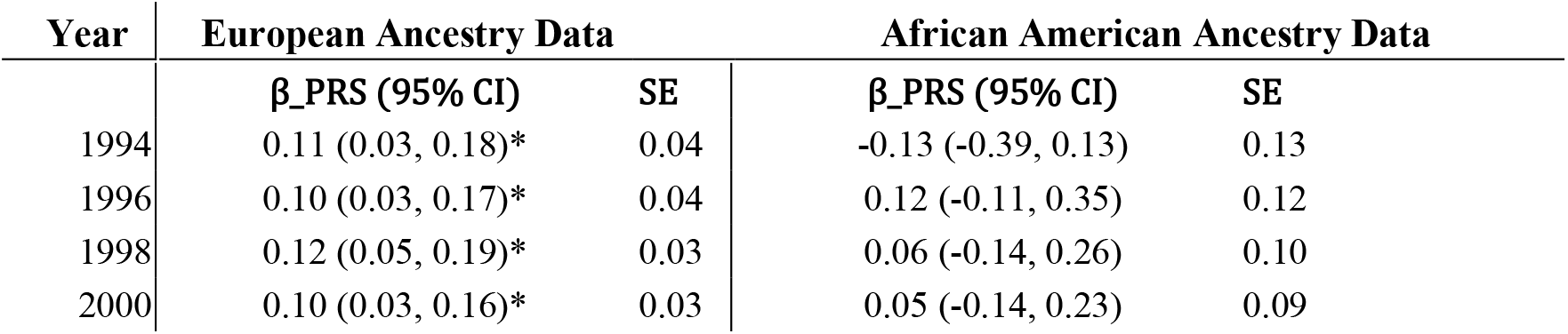
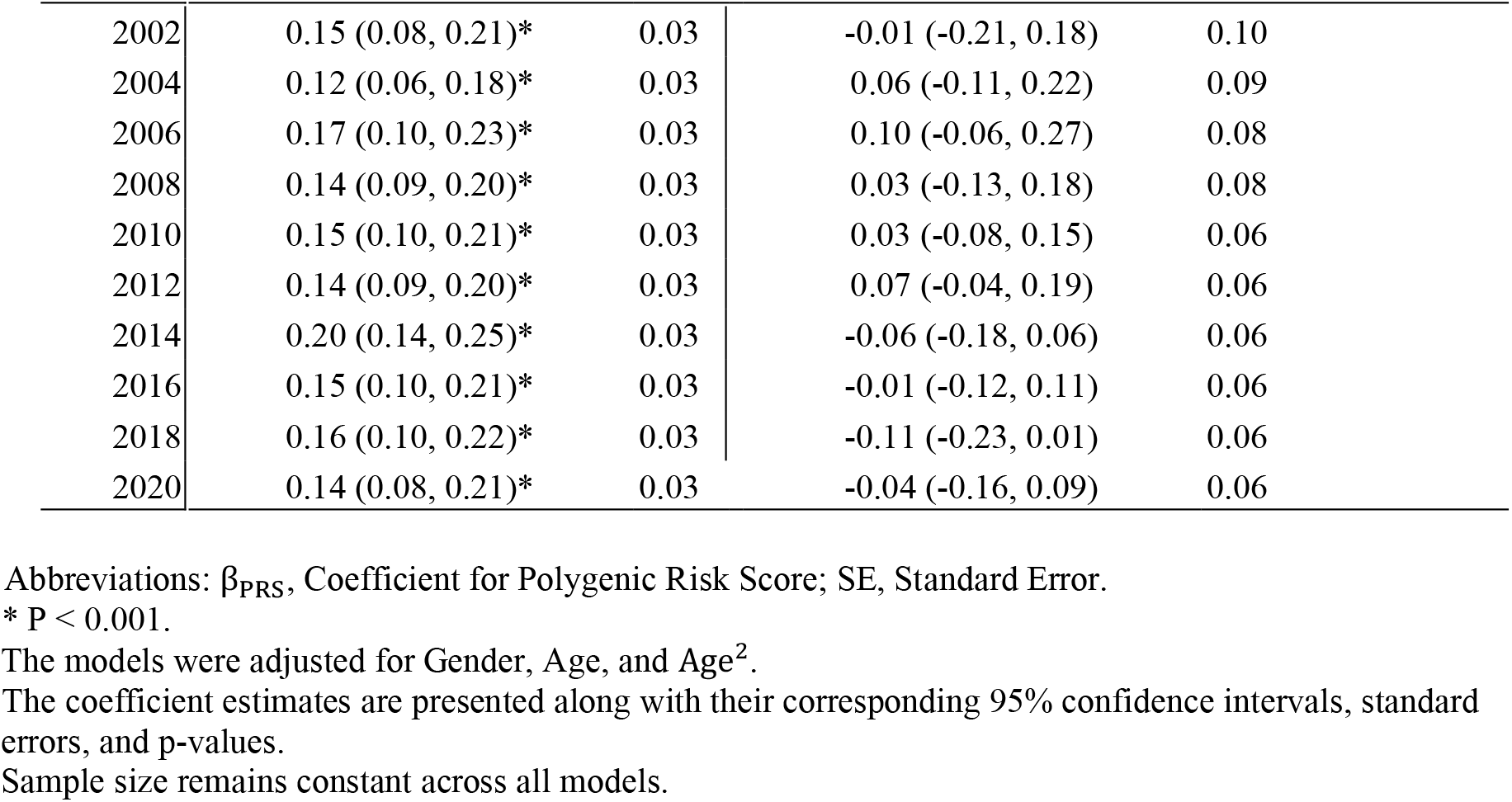
The associations between CESD and PRS (Model 1) for wave years 1994-2020 calculated separately for the White and Black Population Data, Health and Retirement Study Abbreviations: β_PRS_, Coefficient for Polygenic Risk Score; SE, Standard Error. * P < 0.001. The models were adjusted for Gender, Age, and Age^2^. The coefficient estimates are presented along with their corresponding 95% confidence intervals, standard errors, and p-values. Sample size remains constant across all models.

| Year | European Ancestry Data |  | African American Ancestry Data |  |
| --- | --- | --- | --- | --- |
| | $\beta$_PRS (95% CI) | SE | $\beta$_PRS (95% CI) | SE |
| 1994 | 0.11 (0.03, 0.18)* | 0.04 | -0.13 (-0.39, 0.13) | 0.13 |
| 1996 | 0.10 (0.03, 0.17)* | 0.04 | 0.12 (-0.11, 0.35) | 0.12 |
| 1998 | 0.12 (0.05, 0.19)* | 0.03 | 0.06 (-0.14, 0.26) | 0.10 |
| 2000 | 0.10 (0.03, 0.16)* | 0.03 | 0.05 (-0.14, 0.23) | 0.09 |
| 2002 | 0.15 (0.08, 0.21)* | 0.03 | -0.01 (-0.21, 0.18) | 0.10 |
| 2004 | 0.12 (0.06, 0.18)* | 0.03 | 0.06 (-0.11, 0.22) | 0.09 |
| 2006 | 0.17 (0.10, 0.23)* | 0.03 | 0.10 (-0.06, 0.27) | 0.08 |
| 2008 | 0.14 (0.09, 0.20)* | 0.03 | 0.03 (-0.13, 0.18) | 0.08 |
| 2010 | 0.15 (0.10, 0.21)* | 0.03 | 0.03 (-0.08, 0.15) | 0.06 |
| 2012 | 0.14 (0.09, 0.20)* | 0.03 | 0.07 (-0.04, 0.19) | 0.06 |
| 2014 | 0.20 (0.14, 0.25)* | 0.03 | -0.06 (-0.18, 0.06) | 0.06 |
| 2016 | 0.15 (0.10, 0.21)* | 0.03 | -0.01 (-0.12, 0.11) | 0.06 |
| 2018 | 0.16 (0.10, 0.22)* | 0.03 | -0.11 (-0.23, 0.01) | 0.06 |
| 2020 | 0.14 (0.08, 0.21)* | 0.03 | -0.04 (-0.16, 0.09) | 0.06 |

Abbreviations: $\beta_{PRS}$ , Coefficient for Polygenic Risk Score; SE, Standard Error.
\* $P < 0.001$ .

### The associations between CESD, PRS and PAA

Table 3 provides a comprehensive overview of the regression results for White and Black population data, covering Models 2 through 6. The coefficients estimate the impact of various predictor variables on Phenotypic Age Acceleration (PAA).

**Table 3.** The associations between PRS, CESD, and PAA for the White and Black Population Data (Models 2-6), Health and Retirement Study Abbreviations: PRS - Polygenic Risk Score, DS (2016) – Depression Score in 2016, Average DS (1994-2014) – Average Depression Score measured from 1994 to 2014. *** *P* < 0.001, ** *P* < 0.01, * *P* < 0.05. Standard errors are given in parentheses. All models were adjusted for Gender, Age, and Age^2^. Sample size remains constant across all models.

| Variable | White Population |  |  |  |  | Black Population |  |  |  |  |
| --- | --- | --- | --- | --- | --- | --- | --- | --- | --- | --- |
|  | Model 2 | Model 3 | Model 4 | Model 5 | Model 6 | Model 2 | Model 3 | Model 4 | Model 5 | Model 6 |
| PRS | 0.37<br>(0.13)*<br>* |  |  | 0.25<br>(0.13) | 0.20<br>(0.13) | -0.11<br>(0.27) |  |  | -0.10<br>(0.27) | -0.12 (0.27) |
| DS (2016) |  | 0.82<br>(0.08)*** |  | 0.80<br>(0.08)*<br>** |  |  | 0.50<br>(0.13)*<br>** |  | 0.50<br>(0.13)*** |  |
| Average DS (1994-2014) |  |  | 1.21<br>(0.10)*<br>** |  | 1.19<br>(0.10)*** |  |  | 0.78<br>(0.16)*<br>** |  | 1.79<br>(0.16)*** |
| R-squared | 0.06 | 0.08 | 0.09 | 0.08 | 0.09 | 0.01 | 0.02 | 0.03 | 0.02 | 0.03 |
| Adj. R-squared | 0.06 | 0.08 | 0.09 | 0.08 | 0.09 | 0.01 | 0.02 | 0.03 | 0.02 | 0.03 |

\*\*\* $P < 0.001$ , \*\* $P < 0.01$ , \* $P < 0.05$ .

For the White population dataset, Model 2 revealed that the Polygenic Risk Score (PRS) exhibited a statistically significant positive association with PAA (β = 0.37, 95%CI = (0.11, 0.64), *P* < 0.001). However, the inclusion of Depression Score variables (DS in 2016 or Average DS 1994-2014) in subsequent models (3-6) led to the nullification of the PRS coefficient’s significance. Notably, DS in 2016 and Average DS 1994-2014 showed significant positive associations with PAA across Models 3-6 (*P* < 0.001). In Model 3 the results revealed a significant positive association between DS (2016) and PAA (β = 0.82, 95% CI = [0.66, 0.98], *P* < 0.001), indicating that higher depression scores were associated with higher PAA. In Model 4 the analysis revealed a significant positive association between Average DS and PAA (β = 1.21, 95% CI = [1.01, 1.41], *P* < 0.001), indicating that higher average depression scores over the years were associated with higher PAA. In Model 5 the analysis revealed a significant positive association between DS in 2016 and PAA (β = 0.80, 95% CI = [0.64, 0.96], *P* < 0.001), consistent with the findings of Model 3. Finally, in Model 6 the analysis revealed a significant positive association between Average DS and PAA (β = 1.19, 95% CI = [0.99, 1.39], *P* < 0.001), consistent with the findings of Model 4. It is worth mentioning that comparing Models 3 and 4, and Models 5 and 6, the effect of Average DS 1994-2014 on PAA (as captured in Model 4 and 6) is stronger than the effect of DS 2016 (as captured in Model 3 and 5). Gender demonstrated a significant negative association with PAA in all models (*P* < 0.001), suggesting that being male is associated with lower PAA. Age demonstrated a significant negative association with PAA in Models 2,4, and 6. The inclusion of DS variables improved the overall fit of the models, as indicated by the marginal increase in R-squared values.

Conversely, for the Black population dataset, Model 2 revealed a non-significant negative association between PRS and PAA (β = −0.11, *P* = 0.665), which persisted across subsequent models. Model 3 revealed a significant positive association between DS in 2016 and PAA (β = 0.50, 95% CI = [0.34, 0.66], *P* < 0.001). Similarly, in Model 4, a significant positive association was found between Average DS and PAA (β = 0.78, 95% CI = [0.46, 1.10], *P* < 0.001). In Model 5, the analysis also revealed a significant positive association between DS 2016 and PAA (β = 0.50, 95% CI = [0.34, 0.66], *P* < 0.001), consistent with the findings of Model 3. Finally, in Model 6, a significant positive association was observed between Average DS and PAA (β = 1.79, 95% CI = [1.47, 2.11], *P* < 0.001), consistent with the findings of Model 4. Notably, comparing Models 3 and 4, and Models 5 and 6, the effect of Average DS on PAA (as captured in Model 4 and 6) appeared to be stronger than the effect of DS 2016 (as captured in Model 3 and 5). Gender exhibited a significant negative association with PAA in Models 3 through 6 (*P* < 0.001). Age did not show consistent associations across all models. The inclusion of DS variables improved the overall fit of the models, as indicated by the marginal increase in R-squared values.

### Causal Mediation Analysis

Table 4 provides an overview of the mediation analysis results for White and Black population data, respectively. The mediation analysis aimed to explore the relationship between the exposure variable (PRS) and the outcome variable (PAA), mediated through the mediator variable (CESD score at 2016).

**Table 4.** Summary of Causal Mediation Analysis Results for White and Black the Population Data (Wave Year 2016), Health and Retirement Study Abbreviations: ACME, Average Causal Mediation Effect; ADE, Average Direct Effect; CI, Confidence Interval. The analysis included data from 3,806 White and 1,207 Black participants. *P < 0.05.

| Component | White<br>(n=3806) | Population |  | Black<br>(n=1,207) | Population |  |
| --- | --- | --- | --- | --- | --- | --- |
|  | Estimate (95% CI) | Std.<br>Error | P-value | Estimate (95% CI) | Std.<br>Error | P-value |
| ACME | 0.14 (-0.13, 0.43) | 0.14 | 0.284 | -0.003 (-0.07, 0.06) | 0.17 | 0.91 |

| **Component** | **White**<br>**(n=3806)** | **Population** |  | **Black**<br>**(n=1,207)** | **Population** |  |
| --- | --- | --- | --- | --- | --- | --- |
| **ADE** | 0.25 (0.01, 0.50) | 0.12 | 0.046* | -0.09 (-0.63, 0.48) | 0.22 | 0.73 |
| **Total Effect** | 0.26 (0.02, 0.51) | 0.13 | 0.034* | -0.09 (-0.63, 0.48) | 0.23 | 0.71 |
| **Proportion**<br>**Mediated** | 0.12 (0.07, 0.18) | 0.03 | <0.001* | 0.02 (-1.36, 1.43) | 1.07 | 0.88 |

\*P < 0.05.

For the White population, mediation analysis showed no statistically significant effect through CESD scores from 2016 (ACME = 0.144, 95% CI: −0.127 to 0.433, P = 0.284), indicating minimal mediation. The Proportion Mediated was 0.121 (95% CI: 0.069 to 0.180, P < 0.001), suggesting CESD’s limited role in explaining PRS and PAA relationships. For the Black population, mediation analysis found no significant effect through CESD scores from 2016 (ACME = −0.003, 95% CI: −0.07 to 0.06, P = 0.91. The Proportion Mediated was 0.02 (95% CI: −1.36 to 1.43, P = 0.88), confirming CESD’s minimal mediation in PRS and PAA relationships.

## DISCUSSION

This study investigated the relationships between polygenic risk scores, depressive symptoms, and Phenotypic Age Acceleration in a population of individuals of European and African ancestries.

Consistent with prior research ^19–21^, our study identified a significant positive correlation between polygenic risk scores and depression scores in White individuals across all waves. However, among Black individuals, the associations between polygenic risk scores and depression scores tended to be weaker and lacked statistical significance across all waves. Similar patterns were observed in the study by Park et al. ^11^, wherein the efficacy of polygenic risk scores in predicting late-life depression symptoms varied notably based on covariates in African Americans, whereas it consistently predicted such symptoms European American. These highlights potential differences in the genetic architecture underlying depressive symptoms between populations of different ancestries. It should be noted, that the majority of GWAS informing the single nucleotide polymorphisms weights used to calculate polygenic risk scores predominantly involved European ancestry groups^37^. Therefore, the predictive capacity of polygenic risk scores for other ancestry groups, such as African Americans, may not be equivalent ^49^. To address the disparity in predictive capacity across different ancestries, future studies should prioritize the inclusion of diverse ancestral groups in genome-wide association studies (GWAS) to better inform the single nucleotide polymorphisms weights used to calculate polygenic risk scores.

Our research reveals the associations between the polygenic risk score for depression and biological age. A significant positive association was found between polygenic risk score and phenotypic age acceleration, suggesting that individuals with higher polygenic risk scores tend to exhibit greater phenotypic age acceleration. This finding underscores the potential influence of genetic predispositions on biological aging processes. Similarly to the previous analysis of the association between polygenic risk scores and depression scores, the results were significant only for the White population data. In line with the findings of Kuo et al. ^22^, our study underscores the significance of genetic influences on biological aging, particularly regarding PhenoAge. These results reveale distinct genetic determinants of PhenoAge and emphasized the importance of understanding the genetic foundations of age-related processes.

Our findings revealed significant positive associations between depressive symptoms and biological aging, indicating that higher levels of depressive symptoms were associated with accelerated biological aging. This association persisted across different models (for both White and Black population data), including CESD scores measured in 2016 and the average CESD scores spanning from 1994 to 2014. Notably, the effect of average depressive symptoms over time appeared to have a stronger impact on biological aging compared the scores from a single wave year. This suggests that the cumulative burden of depressive symptoms over the years may have a more pronounced effect on biological aging. These results are consistent with the research by Liu et al. ^50^ that shows that depression is associated with faster biological aging.

Including depressive symptoms alongside polygenic risk scores attenuated the significance of the latter, suggesting depressive symptoms may mediate the link between genetic predisposition to depression and biological aging. Multivariate models still showed positive associations between depressive symptoms and biological aging in both White and Black populations. These associations persisted even after adjusting for genetic predisposition, highlighting depressive symptomatology’s significant impact on aging. Addressing depressive symptoms could potentially mitigate accelerated biological aging, promising future research and clinical interventions for healthy aging.

Mediation analysis aimed to explore whether depressive symptoms mediate the relationship between genetic risk and phenotypic age acceleration. No statistically significant mediation effect through depression scores from the wave year 2016 was found for both White and Black population, indicating that depressive symptoms do not significantly mediate the relationship between genetic risk and phenotypic age acceleration.

Several limitations should be considered when interpreting the results of this study. First, the use of self-reported depressive symptoms may introduce bias, and future studies could benefit from including clinical assessments of depression. Additionally, the cross-sectional nature of the data limits our ability to establish causal relationships. Longitudinal studies are needed to better understand the temporal dynamics between genetic risk, depressive symptoms, and biological aging.

Understanding the complex interplay between genetic predisposition, depressive symptoms, and biological aging has important implications for public health and personalized medicine. Our findings underscore the importance of addressing mental health factors, such as depressive symptoms, in efforts to promote healthy aging. Future research should further investigate the underlying mechanisms linking genetic risk, mental health, and biological aging, considering additional environmental and lifestyle factors.

## Data Availability

All data used in the present study are available from the Health and Retirement Study (HRS) upon registration and approval. Genetic data (polygenic scores) require additional application and approval from HRS. Publicly available documentation and procedures for data access can be found at https://hrs.isr.umich.edu/.

## Abbreviations

CESD: Center for Epidemiologic Studies Depression scale
MDD: Major Depressive Disorder
PAA: Phenotypic Age Acceleration
PRS: Polygenic Risk Score
VBS: Venous Blood Study.

